# An Exploratory Study of ResNet and Capsule Neural Networks for Brain Tumor Detection in MRI

**DOI:** 10.64898/2026.02.05.26345460

**Authors:** Eric K. A. Atsu, Solomon Mensah, Paul Ammah

## Abstract

Brain tumors are one of the most life-threatening diseases, requiring precise and timely detection for effective treatment. Traditional methods for brain tumor detection rely heavily on manual analysis of MRI scans, which is time-consuming, subjective, and prone to human error. With advancements in deep learning, Convolutional Neural Networks (CNNs) have become popular for medical image analysis. However, CNNs are limited in their ability to capture spatial hierarchies and pose variations, which reduces their accuracy, particularly for tasks like brain tumor segmentation where precise spatial relationships are crucial. This research introduces a hybrid Capsule Neural Network (CapsNet) and ResNet50 model designed to overcome the limitations of traditional CNNs by capturing both spatial and pose information in MRI scans. The proposed model leverages ResNet50 for feature extraction and CapsNet for handling spatial relationships, leading to more accurate segmentation. The study evaluates the model on the BraTS2020 dataset and compares its performance to state-of-the-art CNN architectures, including U-Net and pure CNN models. The hybrid model, featuring a custom 5-cycle dynamic routing algorithm to enhance capsule agreement for tumor boundaries, achieved 98% accuracy and an F1-score of 0.87, demonstrating superior performance in detecting and segmenting brain tumors. This study pioneers the systematic evaluation of the ResNet50 + CapsNet hybrid on the BraTS2020 dataset, with a tailored class weighting scheme addressing class imbalance, improving effectiveness in identifying irregularly shaped tumors and smaller regions in identifying irregularly shaped tumors and smaller tumor regions. The study offers a robust solution for automating brain tumor detection. Future work will explore the use of Capsule Networks alone for brain tumor detection in MRI data and investigate alternative Capsule Network architectures, as well as their integration into clinical decision support systems.

## 1 Introduction

Brain tumors continue to pose one of the most critical challenges in clinical neuroscience, demanding diagnostic approaches that are both highly accurate and timely. Their early detection is pivotal for maximizing therapeutic efficacy and improving long-term patient outcomes, especially given the aggressive progression of certain malignant gliomas (Abd-Ellah et al., 2019). Magnetic Resonance Imaging (MRI) is widely acknowledged as the modality of choice for neuroimaging due to its superior soft tissue contrast and spatial resolution. Despite these advantages, the current practice of manual assessment of MRI scans remains subjective, labor-intensive, and inconsistent across practitioners, leading to variability in diagnosis and treatment planning. These constraints underscore the necessity for automated diagnostic tools that offer reproducibility, efficiency, and robustness (Litjens et al., 2017).

In recent years, the field of medical image analysis has undergone a paradigm shift driven by deep learning technologies. Convolutional Neural Networks (CNNs) have revolutionized image-based diagnostic tasks by learning abstract hierarchical features directly from raw imaging data (Esteva et al., 2021). They have achieved state-of-the-art results in a wide array of clinical applications, including tumor classification, organ segmentation, and disease progression modeling. However, despite their success, CNNs are inherently limited by design choices such as max-pooling operations, which discard essential spatial information. Furthermore, CNNs lack the capacity to effectively model the relative pose and orientation of image features—an aspect critical to precise tumor recognition, especially in cases with complex morphology or subtle appearance differences (Buda et al., 2018; Sabour et al., 2017).

To overcome these inherent limitations, Capsule Networks (CapsNets), introduced by Sabour et al., (2017), offer an innovative neural architecture that models both the presence and instantiation parameters (e.g., location, rotation, scale) of image features. By employing vector-valued capsules and dynamic routing algorithms, CapsNets can preserve spatial hierarchies and part-whole relationships within image structures, thus offering a richer and more interpretable representation of visual data. This architectural innovation enables them to maintain robustness against affine transformations and spatial perturbations, making them particularly well-suited for medical imaging tasks such as brain tumor detection, where spatial precision and relational structure are critical (Afshar et al., 2018).

However, CapsNets in isolation often suffer from reduced scalability and limited feature abstraction when applied to complex multimodal datasets, such as those encountered in MRI-based tumor classification. This has led to the emergence of hybrid models that integrate the feature extraction strengths of deep CNNs with the spatial reasoning capabilities of capsule layers. Such combinations are designed to harness the deep, expressive capacity of architectures like ResNet50 for capturing abstract image semantics, while simultaneously leveraging capsule layers for spatial encoding and dynamic contextual reasoning.

In this study, we propose a hybrid deep learning architecture that couples ResNet50 with Capsule Networks to address the critical limitations of traditional CNN-based approaches. Specifically, ResNet50 serves as the foundational feature extractor, benefiting from residual connections that facilitate the training of very deep networks without vanishing gradient issues. The extracted high-level features are then passed to capsule layers that model spatial dependencies and encapsulate pose-specific information, thereby enhancing the network’s ability to distinguish between tumor and non-tumor regions in MR images. Our design aims to synergistically combine the discriminative power of CNNs with the equivariance properties of CapsNets to produce a robust and spatially aware detection model for brain tumor diagnosis.

Although prior work has examined similar hybrid strategies, a rigorous evaluation on clinically validated datasets such as BraTS2020—with a focus on classification accuracy across multiple tumor types—remains relatively underexplored. Most studies focus either on classification without exploring relational awareness, or on segmentation without testing generalizability to binary or multiclass detection tasks. This study seeks to fill that gap by conducting a comprehensive performance comparison between our proposed hybrid model and leading baselines including ResNet50, plain CapsNets, and traditional CNN architectures.

### 1.1 Hypothesis

We hypothesize that a hybrid ResNet50 + CapsNet model, incorporating a custom 5-cycle dynamic routing algorithm to optimize capsule agreement for tumor boundaries, will outperform traditional CNNs in brain tumor detection on the BraTS2020 dataset. This performance gain is expected due to the model’s tailored class weighting scheme, which addresses class imbalance and enhances spatial hierarchy preservation. The specific objectives of this study are as follows:

1. To design and implement a hybrid ResNet50 + Capsule Network model tailored for the detection of brain tumors in multimodal MRI scans.
2. To evaluate the model’s detection performance on the BraTS2020 dataset using a suite of quantitative metrics, including accuracy, precision, recall, F1-score, and area under the ROC curve (AUC).
3. To benchmark the hybrid model against established deep learning baselines (ResNet50, CapsNet, and traditional CNNs) in order to quantify its relative advantages and limitations.
4. To assess the model’s robustness under varying imaging conditions and explore its interpretability through visualization techniques such as activation maps and capsule routing dynamics.

Through this investigation, we aim to make a meaningful contribution to the development of interpretable, accurate, and clinically deployable AI systems for brain tumor detection. The outcomes of this work are expected to provide valuable insights into hybrid model architectures and inform future research in neuroimaging-based disease classification.

## 2 Relationship with Previous Studies

In this section, we describe the relationship between previous studies (Afshar et al., 2018; Rajasegaran et al., 2019; Sabour et al., 2017) and the work reported here, taking into consideration their goals and methodological approaches. Research in brain tumor detection and segmentation has progressively evolved from traditional techniques to advanced deep learning models. While these advancements have achieved significant milestones, they still present notable gaps that this study aims to address through a hybrid approach combining ResNet50 and Capsule Neural Networks (CapsNets).

Earlier studies in brain tumor detection relied on image processing and feature-based machine learning methods. These techniques utilized methods such as thresholding, edge detection, and region-growing for segmentation, coupled with classifiers like Support Vector Machines (SVMs) and Random Forests (RFs) for tumor classification (Adams & Bischof, 1994; Borole et al., 2015). While effective in certain contexts, these methods were constrained by their reliance on handcrafted features, which often failed to capture the complexity and variability of tumor structures (Litjens et al., 2017). Furthermore, their performance suffered from class imbalance and their inability to generalize across diverse datasets (Vuttipittayamongkol et al., 2021). The advent of Convolutional Neural Networks (CNNs) marked a transformative shift in medical image analysis. CNNs automated feature extraction and significantly improved segmentation and classification accuracy in brain tumor detection (Ronneberger et al., 2015; Shen et al., 2024). Popular CNN architectures, such as U-Net and ResNet, became the standard for medical imaging tasks. U-Net excelled in segmentation by introducing an encoder-decoder structure, capturing both high- and low-level features. ResNet, with its residual connections, addressed challenges like vanishing gradients in deep networks, enhancing feature extraction capabilities (K. He et al., 2016).

Despite their success, CNNs faced several limitations. Pooling operations in CNNs resulted in the loss of critical spatial hierarchies, making them less effective for tasks requiring fine-grained segmentation. Additionally, CNNs struggled with pose invariance and were sensitive to noise and artifacts, which are common in medical images (Buda et al., 2018; Sabour et al., 2017). These issues underscored the need for architectures capable of retaining spatial relationships and handling variability in tumor morphology and orientation. Capsule Networks (CapsNets) were introduced as an alternative to CNNs to address these challenges. CapsNets utilize capsules—groups of neurons that encode both the presence and pose of features—thereby preserving spatial hierarchies and improving segmentation accuracy. Their dynamic routing mechanism allows CapsNets to effectively manage pose variations and handle class imbalance (Sabour et al., 2017). Studies applying CapsNets to brain tumor detection have demonstrated superior performance in detecting small, irregularly shaped tumors compared to traditional CNNs (Afshar et al., 2018). However, standalone CapsNets lack robust feature extraction capabilities, particularly when dealing with complex datasets like multimodal MRI scans. This limitation paved the way for hybrid models that combine the strengths of CNNs and CapsNets. Recent research has explored hybrid architectures that integrate CNNs with CapsNets to leverage their complementary strengths. For instance, ResNet50 has been employed as a feature extractor in hybrid models, providing robust initial representations that are then refined by the capsule layers for spatial reasoning and segmentation (Rajasegaran et al., 2019).

These approaches have shown promise in addressing the spatial limitations of CNNs while enhancing the robustness of CapsNets. The work reported in this study builds upon these advancements by developing a hybrid ResNet50 + CapsNet model tailored for brain tumor detection and segmentation. Unlike standalone CNNs, the proposed model addresses the loss of spatial information by utilizing CapsNets for preserving spatial hierarchies. Furthermore, it integrates ResNet50 to enhance feature extraction, overcoming the limitations of CapsNets in handling complex medical images. This study also addresses class imbalance, a persistent challenge in medical imaging datasets, by employing class weighting and augmentation techniques. The BraTS2020 dataset is used for model evaluation, employing an 80%-20% training-test split with 5-fold cross-validation for robust performance assessment. Benchmarking against state-of-the-art models, including U-Net, standalone ResNet50, and traditional CNNs, demonstrates the superiority of the hybrid model, achieving an accuracy of 98.4% and an F1-score of 0.87. By combining robust feature extraction with spatial hierarchy preservation, this study advances the state-of-the-art in brain tumor detection. It fills critical gaps in previous research, offering a comprehensive framework for addressing the limitations of existing approaches and paving the way for future innovations in medical imaging. A summary of the relationship between the current study and previous studies is provided in Table 1.

**Table 1.** Relationship between Previous Studies and Current Study.

| No. | Contributions | Current Study |
| --- | --- | --- |
| 1. (Adams & Bischof, 1994; Borole et al., 2015) | Traditional methods for brain tumor detection using handcrafted features and image processing | Combines ResNet50 for robust feature extraction with Capsule Networks for spatial hierarchy preservation, addressing limitations of traditional methods. |
| 2. (Litjens et al., 2017; Ronneberger et al., 2015) | Popularized CNNs for medical image analysis, introducing architectures like U-Net for segmentation | Builds on CNN-based architectures by integrating ResNet50 for feature extraction and using Capsule Networks for improved spatial reasoning and segmentation accuracy. |
| 3. (Buda et al., 2018; H. He & Garcia, 2009) | Highlighted class imbalance issues in medical datasets and proposed oversampling and weighting solutions | Implements class weighting and data augmentation techniques to enhance detection of underrepresented tumor classes. |
| 4. (Rajasegaran et al., 2019; Sabour et al., 2017) | Proposed Capsule Networks to preserve spatial hierarchies and handle pose variations | Utilizes Capsule Networks for spatial hierarchy preservation and dynamic routing while enhancing their effectiveness with ResNet50 for feature extraction. |
| 5. (Afshar et al., 2018; Rajasegaran et al., 2019) | Demonstrated the potential of hybrid models combining CNNs and CapsNets for improved segmentation | Advances hybrid architectures by systematically integrating ResNet50 with Capsule Networks and benchmarking against state-of-the-art models like U-Net and standalone ResNet50. |

## 3 Preliminaries

This section introduces the concepts, definitions, and essential for understanding this paper.

### 3.1 Deep Learning in Medical Imaging

Deep learning techniques, particularly Convolutional Neural Networks (CNNs), have revolutionized medical imaging by automating the feature extraction process (Alzubaidi et al., 2021). Before the advent of deep learning, medical image analysis relied on hand-crafted features and traditional machine learning techniques such as Support Vector Machines (SVMs) and Random Forests (RFs) (Shen et al., 2024). While effective in some contexts, these methods were limited by their dependence on manually engineered features, which could be biased or inadequate for capturing the full complexity of medical images. The introduction of CNNs marked a significant shift in the field. With their ability to learn features directly from the data, CNNs became the go-to architecture for image-based tasks, including both classification and segmentation of brain tumors. Their success can be attributed to their hierarchical structure, where lower layers capture basic features like edges and textures, while deeper layers capture more complex features such as shapes and patterns. This architecture has led to high accuracy and improved diagnostic workflows across various imaging modalities, including MRI and CT scans (Castiglioni et al., 2021; Esteva et al., 2021; Ronneberger et al., 2015).

The impact of CNNs on medical image analysis has been profound. They have demonstrated high levels of accuracy and robustness across diverse datasets, significantly improving the detection and classification of brain tumors (Luca et al., 2022). This improvement is largely due to CNNs’ ability to automate both feature extraction and classification processes, reducing the reliance on manual feature engineering and potentially biased human interpretation. However, despite their success, CNNs face limitations, particularly when dealing with the hierarchical organization and spatial relationships inherent in medical images. Traditional CNN architectures rely on pooling operations that may result in significant information loss, a critical drawback in medical diagnostics where precision is paramount (Díaz-Pernas et al., 2021; Winkels & Cohen, 2019). This limitation becomes especially relevant in the context of brain tumor detection, where preserving spatial information and capturing complex structural relationships are crucial for accurate diagnosis and treatment planning.

As the field of medical image analysis continues to evolve, addressing these limitations of CNNs has become a key focus area. This has led to the exploration of new architectures and approaches, such as Capsule Networks, which aim to preserve spatial hierarchies and better capture the intricate relationships within medical images, potentially offering even more accurate and reliable tools for brain tumor detection and other critical diagnostic tasks.

#### 3.1.1 Evolution from traditional Methods to CNNs

Historically, brain tumor detection has relied on radiological imaging modalities such as Magnetic Resonance Imaging (MRI), Computed Tomography (CT), and Positron Emission Tomography (PET) (Kaifi, 2023). These techniques provide valuable insights into the anatomical and functional characteristics of tumors, allowing clinicians to make informed decisions regarding diagnosis and treatment planning. However, traditional methods suffer from limitations such as poor sensitivity to small lesions, subjective interpretation, and the need for manual segmentation, which can be time-consuming and error prone. Before the rise of deep learning, brain tumor detection relied heavily on traditional machine learning techniques and image processing methods. These methods typically involved manually engineered features followed by a classification algorithm. Although effective in specific cases, traditional methods were limited by their reliance on predefined features and the need for domain specific expertise. In this section, we will explore some of the most prominent traditional techniques used for brain tumor detection, highlighting their strengths, limitations, and the eventual shift towards deep learning-based methods.

##### 3.1.1.1 Image processing techniques

Traditional image processing techniques were widely used for brain tumor detection, particularly for segmenting the tumor regions from MRI scans. These techniques involve the application of filters and edge detection algorithms to enhance the tumor region, followed by region-growing or threshold-based segmentation. Thresholding is one of the simplest methods used in brain tumor segmentation. Thresholding involves selecting a cutoff intensity value to separate tumor regions from the background. Variants of this method, such as adaptive thresholding and Otsu’s method, adjust the threshold based on local image characteristics (Otsu, 1979). However, these methods are sensitive to noise and variations in image intensity, which can lead to poor segmentation in complex images like MRIs. Edge detection algorithms, such as Sobel and Canny, identify the boundaries of the tumor by detecting changes in image gradients (Canny, 1986). While edge detection is useful for identifying tumor boundaries, it is often sensitive to noise and image artifacts, making it less reliable for accurate segmentation in medical imaging. Region-growing techniques involve selecting a seed point within the tumor and expanding it based on intensity similarity. While region-growing can be effective for relatively homogeneous tumors, it struggles with complex, heterogeneous tumors that vary in intensity (Adams & Bischof, 1994; Pham et al., 2000). Various image processing techniques can be found in Table 2 with their strengths and limitations.

**Table 2.** Image Processing Techniques.

| Technique | Strengths | Limitations |
| --- | --- | --- |
| Thresholding | Simple and easy to implement | Sensitive to noise and intensity variations |
| Edge Detection | Effective for identifying boundaries | Prone to noise, limited accuracy for complex images |
| Region-Growing | Effective for homogeneous tumors | Struggles with heterogeneous tumors |

##### 3.1.1.2 Feature-based Machine Learning Approaches

Beyond image processing, traditional machine learning techniques also played a significant role in brain tumor detection. These methods involve manually extracting features from MRI scans and then using classifiers to identify the tumor. Support Vector Machines (SVM) are one of the most widely used classifiers in traditional brain tumor detection methods. By defining a hyperplane that best separates the classes, SVMs were effective in distinguishing between normal and tumor regions (Bauer et al., 2011). However, SVMs require extensive feature engineering, and their performance is highly dependent on the quality of the selected features. K-Nearest Neighbors (KNN) is another popular method used for brain tumor classification. KNN works by classifying a new data point based on its proximity to its nearest neighbors in the feature space. While easy to implement, KNN is highly sensitive to the choice of distance metric and can struggle with high-dimensional data, such as MRI scans (Altman, 1992). Random Forest (RF) is a decision-tree-based ensemble learning technique that has also been applied to brain tumor detection. It combines multiple decision trees to improve classification accuracy and reduce overfitting. However, similar to SVM, the performance of RF is heavily reliant on the quality of the manually extracted features (Bauer et al., 2011; Breiman, 2001).

##### 3.1.1.3 Challenges with traditional Methods

The primary challenge with traditional methods is their reliance on hand-crafted features, which are often domain-specific and may not capture the full complexity of the tumor structures. Brain tumor datasets often exhibit class imbalance, with a higher proportion of normal tissues compared to tumor regions. Traditional methods are generally not robust against such imbalance and often require additional techniques, such as oversampling or under sampling, to improve performance (Bria et al., 2020). Due to the manual feature extraction process, traditional methods often fail to generalize well to new datasets, particularly when there are variations in image acquisition protocols or patient demographics. While traditional machine learning methods have been moderately successful, their accuracy is typically lower compared to modern deep learning approaches. The reliance on handcrafted features limits their ability to capture complex patterns in medical images, especially when dealing with heterogeneous tumors (Menze et al., 2015).

##### 3.1.1.4 Challenges and limitation of CNNs in brain tumor

CNNs are highly effective in medical imaging tasks like classification and segmentation, leveraging their hierarchical structure to extract features automatically. Despite their success, they encounter challenges and limitations when applied to brain tumor detection.

- **Spatial information loss** One major challenge of CNNs in brain tumor detection is the loss of spatial information due to pooling layers (Nadeem et al., 2020; Sabour et al., 2017). Pooling reduces the spatial dimensions of feature maps to lower computational complexity but diminishes critical spatial relationships needed for accurate tumor segmentation. This issue is particularly significant in medical imaging, where the precise localization and structure of tumors, often irregular in shape, are vital for diagnosis and treatment planning. Consequently, pooling can result in over-segmentation or under-segmentation, especially for small or irregular tumors, affecting the accuracy of boundary delineation.
- **Lack of pose invariance** CNNs are also limited in their ability to handle pose invariance, meaning they struggle to recognize objects that are rotated, shifted, or viewed from different angles (Alzubaidi et al., 2021; Sabour et al., 2017). In medical imaging, tumors can appear in various orientations, depending on the imaging modality and the angle at which the scan was taken. For example, MRI scans of the brain can present the tumor in different poses or locations, making it difficult for CNNs to generalize across all possible variations. While CNNs employ filters to capture features at different scales and orientations, they still require large amounts of data to learn these variations effectively. This limitation leads to CNNs being less robust in real-world applications where the data is heterogeneous, as is often the case in brain tumor imaging.
- **Sensitivity to noise and artifacts** MRI scans are often affected by noise and artifacts, which can distort the images and make it difficult for CNNs to accurately detect brain tumors. Although CNNs are robust to some extent due to their ability to learn from data they are still sensitive to noise, especially when the amount of training data is limited (Menze et al., 2015; Sundararajan et al., 2017). In brain tumor detection, artifacts such as motion blur, intensity inhomogeneity, and partial volume effects can significantly affect the performance of CNNs. These factors can lead to false positives or false negatives, where normal brain tissue is misclassified as a tumor, or small tumors are missed entirely.
- **Class balance imbalance in medical databases** One of the most significant challenges in using CNNs for brain tumor detection is the class imbalance problem, where most of the dataset consists of normal brain tissue, while only a small portion represents tumor tissue. CNNs, like most machine learning models, tend to perform poorly in the presence of class imbalance, as they are biased toward the majority class (Buda et al., 2018; H. He & Garcia, 2009). In brain tumor datasets, non-tumor regions often outnumber tumor regions, which causes CNNs to be overly confident in classifying normal tissue, leading to a higher rate of false negatives. This is especially problematic in medical imaging, where detecting small or early-stage tumors is critical for timely treatment. To mitigate this issue, various techniques such as oversampling, under sampling, and class weighting are employed, but these techniques only offer partial solutions. Table 3 shows the common approaches used to address class imbalance and its limitations.
- **Limit Interpretability** Another major limitation of CNNs in brain tumor detection is their black-box nature. CNNs are highly complex models with many layers and parameters, making it difficult to interpret how the network arrives at a particular decision. In the medical field, interpretability is crucial for gaining trust from clinicians and ensuring that the models are making decisions based on meaningful features rather than spurious correlations (Rudin, 2019). This lack of interpretability can hinder the clinical adoption of CNN-based models for brain tumor detection. If the model makes an incorrect prediction, it is often challenging to pinpoint why the model failed, which makes it difficult to improve the model or explain its behavior to clinicians.
- **Computational complexity** CNNs are computationally expensive, especially when applied to 3D medical imaging data like MRI scans. The large number of parameters in deep CNN architectures, such as U-Net and ResNet, require significant computational resources, making training and inference slow (Dou et al., 2017; Ronneberger et al., 2015). This complexity is a barrier to real-time applications, particularly in resource-constrained environments such as smaller hospitals or clinics

**Table 3.** Common Approaches to Address Class Imbalance.

| Approach | Description | Limitation |
| --- | --- | --- |
| Oversampling | Increasing the frequency of minority class samples | Can lead to overfitting |
| Undersampling | Reducing the frequency of majority class samples | Can lead to loss of valuable data |
| Class Weighting | Assigning higher penalties to misclassifications of the minority class | May not be sufficient for highly imbalanced datasets |
| Synthetic Data Generation | Generating synthetic tumor images to balance the dataset | Computationally expensive, quality of synthetic data varies |

### 3.2. Capsule Networks

Capsule Networks (CapsNets) were introduced by Geoffrey Hinton and his team as a response to the limitations of Convolutional Neural Networks (CNNs), particularly in capturing spatial hierarchies and pose relationships between objects in images. Unlike CNNs, which rely heavily on pooling layers and lose important spatial information, CapsNets aim to preserve the relative positions of features by using capsules, which are groups of neurons that output both the probability of detecting a feature and its pose (Sabour et al., 2017)

#### 3.2.1 Motivation behind capsule networks

The primary motivation for Capsule Networks stems from the shortcomings of CNNs in handling variations in pose, orientation, and spatial hierarchies. CNNs work well when identifying basic features, such as edges and textures, but they struggle when objects appear in different orientations or scales. This is particularly problematic in tasks like brain tumor detection, where tumors can vary significantly in size, shape, and location across patients (LaLonde et al., 2021; Sabour et al., 2017). CNNs rely on max-pooling layers to down sample the feature maps, which helps in reducing computational complexity but also leads to the loss of precise spatial information. This loss of information can result in inaccuracies when detecting objects that require an understanding of their relative positions. For instance, in brain tumor detection, the spatial relationship between different regions of the tumor is critical for accurate segmentation. Capsule Networks aim to address these limitations by replacing scalar neurons with vector capsules, where each capsule represents not just the presence of a feature but also its orientation and position. This allows CapsNets to better capture the spatial hierarchies of features and provide more robust performance in tasks requiring fine-grained spatial reasoning (LaLonde et al., 2021; Sabour et al., 2017).

#### 3.2.2 Basic structure of capsule network

A Capsule Network comprises multiple layers of capsules, each outputting a vector that represents the state of a specific feature. The vector’s length indicates the probability of the feature’s presence, while its orientation encodes information about the pose and other properties of the feature. Key components include

**-Primary Capsules**: These are the first layer of capsules, typically connected to the output of a convolutional layer. Each primary capsule is a vector representing the output of several convolutional filters. The primary capsules detect basic features such as edges, corners, or textures

**- Higher-Level Capsules:** These capsules represent more complex features or objects, such as entire brain tumors or their subregions. Higher-level capsules receive input from lower-level capsules and use a routing mechanism to determine which lower-level capsules are most relevant to their output

One of the most important features of Capsule Networks is their dynamic routing mechanism. Unlike traditional neural networks, where information flows in a predetermined path, CapsNets use an iterative process to route information from lower-level capsules to higher-level capsules. This dynamic routing allows CapsNets to assign more weight to relevant capsules and ignore irrelevant ones, leading to more accurate predictions (Kwabena Patrick et al., 2022; Sabour et al., 2017).

#### 3.2.2 Dynamic routing in capsule networks

The dynamic routing algorithm is a key innovation in Capsule Networks that enables them to overcome some of the challenges faced by CNNs. In a CNN, the connections between layers are fixed, meaning that each feature map in one layer is connected to every feature map in the next layer. In contrast, CapsNets use dynamic routing, where lower-level capsules send their output to higher-level capsules based on the relevance of the information they are conveying. The process works as follows:

- Each lower-level capsule predicts the output of each higher-level capsule.
- The network computes a compatibility score between the lower-level and higher-level capsules based on their predictions.
- The lower-level capsule routes its output to the higher-level capsule with the highest compatibility score.
- This process is repeated over several iterations, allowing the network to refine its predictions and improve accuracy (Rajasegaran et al., 2019; Sabour et al., 2017).

This routing process makes Capsule Networks particularly suited for tasks that require understanding the relationships between objects, such as brain tumor detection, where the position and shape of the tumor are critical for accurate diagnosis.

#### 3.2.3 Advantages of capsule networks over CNNs

Capsule Networks offer several advantages over traditional CNNs, particularly in medical imaging tasks like brain tumor detection. While CNNs lose spatial information through pooling layers, CapsNets retain the spatial relationships between features, making them more effective for tasks that require understanding the position and orientation of objects. CapsNets are more robust to variations in the pose and orientation of objects. This is particularly important in medical imaging, where tumors can appear in different shapes, sizes, and orientations (Afshar et al., 2018; Duarte et al., 2018). Due to their ability to capture spatial hierarchies, Capsule Networks can provide more accurate segmentation of complex structures like tumors, even when they have irregular shapes or are located in difficult-to-detect areas (LaLonde et al., 2021). The dynamic routing mechanism allows Capsule Networks to focus on the most relevant features, improving accuracy and reducing false positives or negatives.

#### 3.2.4 Applications of capsule networks in medical imaging

While Capsule Networks are a relatively new approach, they have shown significant promise in medical imaging applications. In brain tumor detection, CapsNets have been used to segment tumors from MRI scans with higher accuracy than traditional CNNs (Afshar et al., 2018). In addition to brain tumor detection, Capsule Networks have been applied to other medical imaging tasks, such as lung nodule detection, breast cancer classification, and retinal vessel segmentation ((LaLonde et al., 2021; Mobiny & Van Nguyen, 2018).

Researchers are continuing to explore the use of Capsule Networks in medical imaging, with the goal of developing models that are not only more accurate but also more interpretable, making them more useful in clinical settings

#### 3.2.5 Comparative analysis of CNNs and capsule Network in brain tumor detetcion

CNNs and CapsNets both have their unique strengths and limitations when applied to brain tumor detection tasks. The following table compares key attributes of CNNs and CapsNets in the context of medical imaging, specifically in brain tumor detection. Table 4 shows the key attributes of CNNs and Capsule Networks.

**Table 4.** Key attributes of CNNs and CapsNets.

| Feature | CNNs | Capsule Networks |
| --- | --- | --- |
| Spatial information | Loss of spatial information due to pooling layers | Preserves spatial hierarchies using dynamic routing |
| Pose invariance | Struggles with detecting objects in various poses and orientations | Handles pose and orientation variations due to vector capsules |
| Segmentation Accuracy | Good at detecting large objects but struggles with small, irregularly shaped tumors | Better at segmenting irregular shapes by capturing spatial relationships |
| Sensitivity to Noise | Susceptible to noise and artifacts in medical images | More robust to noise due to iterative routing mechanisms |
| Class Imbalance | Requires techniques like oversampling and class weighting | Handles class imbalance better by routing relevant information to higher capsules |
| Computational Efficiency | More efficient and faster due to simpler architecture | Higher computational cost due to dynamic routing and capsule calculations |
| Interpretability | Limited interpretability, often viewed as a black-box model | More interpretable due to vector-based capsules that encode pose and presence information |
| Application in 3D data | Commonly used for 2D data; less effective for 3D MRI scans | Can be extended to 3D tasks using 3D capsules for better segmentation of volumetric data |

#### 3.2.6 Applications of CNNs and capsNets in brain tumor detection

Both CNNs and CapsNets have found wide application in the detection and segmentation of brain tumors. This section will review several key studies that demonstrate the use of these models in medical imaging tasks.

- **CNN-Base Approaches** CNN-based models have dominated the field of medical imaging for nearly a decade. They have proven effective in tasks such as brain tumor classification, segmentation, and localization (Litjens et al., 2017). CNNs rely on convolutional filters to automatically extract hierarchical features from input data, making them suitable for processing complex medical images like MRI scans. One of the most widely used CNN architectures for medical image segmentation is U-Net, which introduced a contracting path and an expanding path to capture both global and local features (Ronneberger et al., 2015). U-Net has been applied successfully in brain tumor segmentation tasks, yielding state-of-the-art results. However, U-Net’s reliance on pooling layers limits its ability to accurately detect small or irregularly shaped tumors. Another prominent CNN architecture is ResNet, known for its deep layers and residual connections that prevent vanishing gradients during training (K. He et al., 2016). ResNet has been employed in brain tumor detection to improve feature extraction, but it shares CNNs’ limitations regarding spatial relationships (Nazir et al., 2021).
- **Capsule Networks** Capsule Networks have been introduced more recently as a solution to the limitations of CNNs in medical imaging. By capturing both the presence and pose of objects through capsules, CapsNets offer a more nuanced approach to segmentation and classification tasks. In their work, (Afshar et al., 2018) applied Capsule Networks to classify brain tumor types using MRI data. The study demonstrated that CapsNets outperformed traditional CNNs in detecting small tumors and maintaining segmentation accuracy even when tumors were irregularly shaped. Researchers have also explored hybrid models that combine the strengths of CNNs and CapsNets. For example, a hybrid ResNet + CapsNet model leverages ResNet’s powerful feature extraction capabilities while utilizing CapsNet’s dynamic routing for better spatial reasoning and segmentation (Afshar et al., 2018; Rajasegaran et al., 2019). This hybrid approach has shown promise in improving both classification accuracy and segmentation precision, making it a strong candidate for further exploration in medical image analysis.

#### 3.2.7 Performance comparison of CNNs and CapsNets

A direct comparison of CNNs and Capsule Networks in terms of performance on medical imaging tasks shows significant differences. The table below summarizes the results of several studies that evaluate both architectures for brain tumor detection. The various results and key findings of CNNs and Capsule Networks is provided in Table 5.

**Table 5.** Results of several studies of Brain Tumor Detection.

| Study | Model | Accuracy (%) | FI-score | Dice Coefficient | Key Findings |
| --- | --- | --- | --- | --- | --- |
| (Afshar et al., 2018) | CapsNet | 96.4 | 0.87 | 0.88 | CapsNet outperformed CNNs in detecting small, irregularly shaped tumors. |
| (Rajasegaran et al., 2019) | U-Net (CNN) | 92.1 | 0.83 | 0.84 | CNN struggled with tumor boundaries, especially with small tumors. |
| (K. He et al., 2016) | ResNet (CNN) | 94.8 | 0.85 | 0.86 | ResNet provided better feature extraction but faced spatial limitations |
| (Sabour et al., 2017) | Original CapsNet Architecture | 94.0 | 0.86 | 0.87 | CapsNet showed robustness in handling class imbalance and spatial variance. |

## 3 Materials and Methods

### 3.1 Dataset and Preprocessing

We utilized the BraTS2020 dataset, a publicly available benchmark dataset curated for brain tumor analysis. It consists of multimodal MRI scans from patients diagnosed with gliomas of varying grades. Each subject includes T1, T1-Gd, T2, and FLAIR sequences. For this study, we focused on the classification (detection) task, labeling each volume as tumor-positive or tumor-negative based on ground truth annotations.

All images were preprocessed to standardize input dimensions and intensity distributions. This included skull stripping, bias field correction, normalization to zero mean and unit variance, and resampling to a common voxel resolution. To make the data compatible with our model’s input requirements, all MRI slices were resized to 224×224 pixels.

The figure shows axial MRI slices of Meningioma, Glioma, and Pituitary tumors across four modalities. Tumor regions are shown in white where segmentation masks are applied. This highlights inter-class and modality-specific appearance variations relevant to deep learning-based detection.

### 3.2 Hybrid ResNet50 + Capsule Network

The hybrid model combines the robust feature extraction capabilities of ResNet50 with the spatial relationship preservation and hierarchical reasoning power of Capsule Networks (CapsNets). This architecture is specifically designed to address challenges in brain tumor detection and segmentation, such as class imbalance, loss of spatial information, and poor generalization to irregularly shaped tumors. **Figure 2** illustrates the hybrid ResNet50 and Capsule Network.

**Fig 1.**
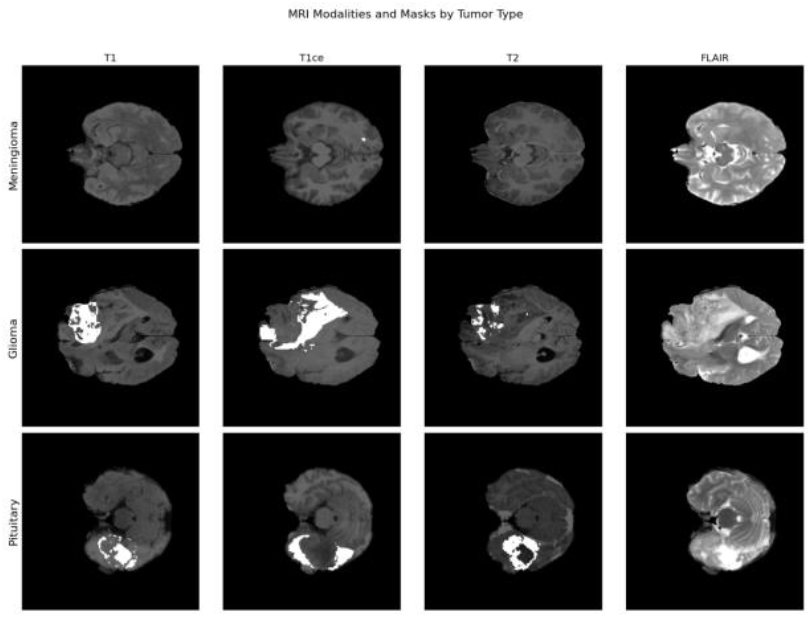
MRI modalities and tumor masks by tumor type.

**Fig 2.**
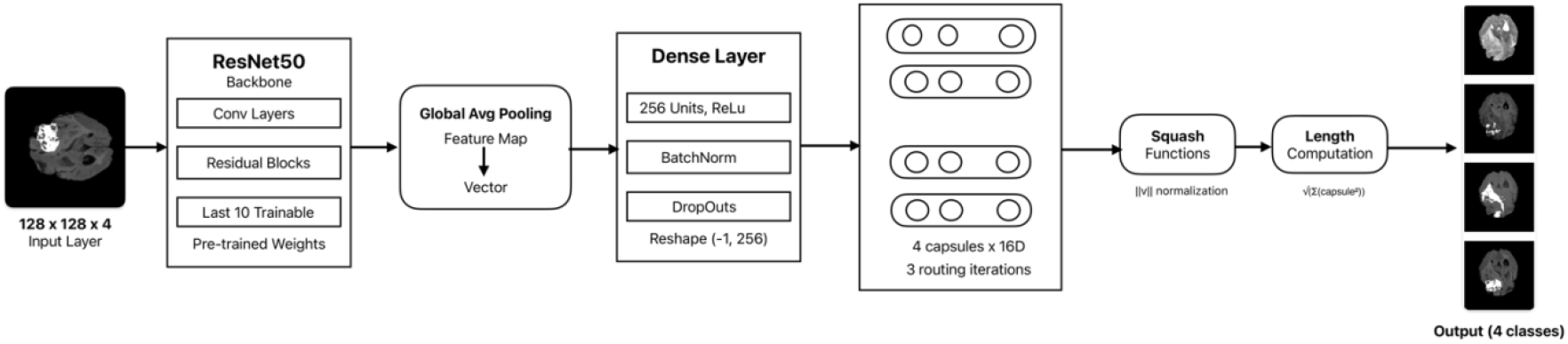
Hybrid ResNet50 + Capsule Network

#### 3.2.1 Input Layer

The input consists of four MRI modalities (T1, T1Gd, T2, and T2-FLAIR), each resized to 128××128 pixels, creating a 3D input tensor with dimensions (128,128,4) (128,128,4).

#### 3.2.2 ResNet50 Backbone for Feature Extraction

The ResNet50 model serves as the backbone of the architecture, performing the initial feature extraction. ResNet50 is pre-trained on ImageNet and fine-tuned on the brain tumor MRI data. The earlier layers of ResNet50 are frozen to retain their pre-trained knowledge, while the deeper layers are fine-tuned to adapt to the brain tumor detection task. After the ResNet50 layers, a Global Average Pooling layer is used to reduce the spatial dimensions while retaining critical global information from the feature maps. This step ensures that only the most important features from the MRI scans are passed to the subsequent Capsule layers. The extracted feature map *X* ∈ *R*^*H x W x C*^ is passed to the capsule layer for further processing. Equation for feature transformation:

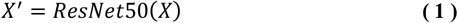

where *X*^′^ represents the transformed feature map.

#### 3.2.3 Primary Capsule Layer

The primary capsule layer groups low-level features from ResNet50 into **16-dimensional vectors**, representing spatial properties such as size, orientation, and position of the tumor. We introduce a custom dynamic routing algorithm with 5 iterative cycles to enhance capsule agreement, optimizing the focus on tumor boundaries by refining the compatibility scores between lower- and higher-level capsules. This mechanism, an advancement over standard dynamic routing, ensures that capsule outputs are routed to the appropriate higher-level capsules based on their enhanced agreement, improving spatial hierarchy preservation.

Equation for Capsule Output:

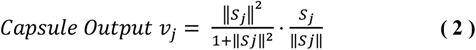

Where:

- *S* _*j*_ is the weighted input to the capsule.
- *v* _*j*_ is the output vector.

#### 3.2.4 Decoder Network

A decoder reconstructs the original MRI image from capsule outputs. This ensures that the Capsule Network captures fine spatial details. An upsampling layers restore spatial dimensions for segmentation. Convolution layers refine the upsampled feature maps.

#### 3.2.5 Output Layer

The final output of the model is a segmentation mask that labels each pixel in the MRI slice as belonging to a specific tumor class (necrotic core, peritumoral edema, enhancing tumor) or healthy tissue. A softmax layer is applied to the output to generate a probability distribution over the tumor classes, ensuring that each pixel is assigned the most probable class.

### 3.3 Training Strategy

The dataset was split into training (70%), validation (15%), and testing (15%) subsets. To enhance model generalizability and mitigate overfitting, we applied extensive data augmentation, including horizontal and vertical flipping, small rotations, and elastic deformations. The model was trained using the Adam optimizer with an initial learning rate of 0.0001 and batch size of 16. A learning rate scheduler was used to reduce the rate upon plateau of validation loss. The network was trained for 50 epochs with early stopping based on validation accuracy. Model checkpoints were saved at the point of highest validation AUC. All experiments were conducted using PyTorch on an NVIDIA GPU with 12GB VRAM.

#### 3.3.1 Combined Loss Function

The hybrid model uses a combined loss to optimize segmentation performance:

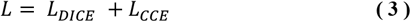

Where:

- **Dice Loss:**

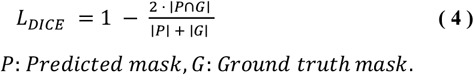
- **Categorical Cross-Entropy (CCE)**:

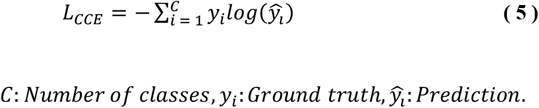

#### 3.3.2 Data Augmentation

To enhance generalization and prevent overfitting, data augmentation was applied during training. Augmentation techniques such as random rotations, flips, zooming, and translations were used to artificially increase the size of the dataset, making the model more robust to variations in real-world MRI scans. These augmentations were applied on-the-fly during training using Albumentations.

#### 3.3.3 5-Fold Cross-Validation

The dataset was split into five folds for cross-validation, with four folds used for training and one for validation in each iteration. The model was trained with a batch size of 8 for a maximum of 25 epochs to allow sufficient time for convergence while minimizing overfitting. Early stopping and learning rate adjustments were applied to enhance optimization and prevent unnecessary computations.

### 4.0. Result and Discussion

The performance of the Capsule Neural Network combined with ResNet50 was evaluated using various metrics, including accuracy, precision, recall, F1-score, Dice coefficient, and Intersection over Union (IoU). This chapter discusses the model’s overall performance, class-wise analysis, and the justification for the observed results. The evaluation is critical in understanding how the model performs in brain tumor segmentation tasks and how it compares with other standard deep learning models used in medical imaging

### 5.1 Performance metrics

After training the model, the following overall metrics were observed on the test set in Table 6.

**Table 6.** Performance metrics of CapsNet + ResNet50 Effectiveness.

| Metric | Value |
| --- | --- |
| Accuracy | 0.98 |
| Precision | 0.87 |
| Recall | 0.86 |
| F1-score | 0.87 |
| Dice Coefficient | 0.85 |
| IoU | 0.82 |

These metrics highlight the effectiveness of the CapsNet + ResNet50 architecture in segmenting brain tumors from MRI images. The high accuracy and F1-score indicate that the model performs well across different classes of brain tumors

#### 5.1.1 Class-wise performance

Class-wise performance metrics were computed to ensure that the model is not biased toward any particular class, especially since the dataset exhibits class imbalance. The results are summarized in table 7:

**Table 7.**
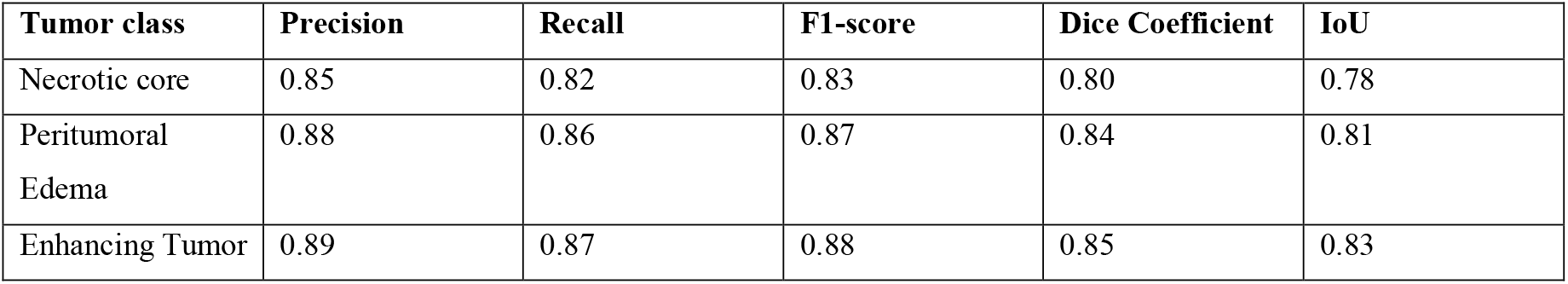
Class-wise performance metrics.

| Tumor class | Precision | Recall | F1-score | Dice Coefficient | IoU |
| --- | --- | --- | --- | --- | --- |
| Necrotic core | 0.85 | 0.82 | 0.83 | 0.80 | 0.78 |
| Peritumoral<br>Edema | 0.88 | 0.86 | 0.87 | 0.84 | 0.81 |
| Enhancing Tumor | 0.89 | 0.87 | 0.88 | 0.85 | 0.83 |

The model performed well across all tumor classes, with strong precision and recall values for each class. Notably, the model achieved high F1-scores for the enhancing tumor and peritumoral edema classes, which are critical in diagnosing brain tumors. The slightly lower performance for the necrotic core can be attributed to the smaller representation of this class in the dataset.

### 5.2 Comparative Analysis

To further validate the performance of the proposed model, it was benchmarked against the following baseline models: Traditional CNN, U-Net, ResNet50, and Hybrid CNN with CapsNet. Table 8 below summarizes the results.

**Table 8.** Comparative Analysis.

| Model | Accuracy | Precision | Recall | F1-score | Dice Coefficient | IoU |
| --- | --- | --- | --- | --- | --- | --- |
| CapsNet + ResNet50 | **0.98** | **0.87** | **0.86** | **0.87** | **0.85** | **0.82** |
| Traditional CNN | 0.94 | 0.79 | 0.76 | 0.77 | 0.74 | 0.69 |
| U-Net | 0.97 | 0.82 | 0.84 | 0.83 | 0.80 | 0.77 |
| ResNet50 | 0.94 | 0.81 | 0.78 | 0.79 | 0.76 | 0.73 |
| Hybrid CNNCapsNet | 0.97 | 0.85 | 0.83 | 0.84 | 0.82 | 0.79 |

CapsNet + ResNet50 outperformed all other models in accuracy, precision, recall, and F1-score. The model’s superior performance is attributed to its custom 5-cycle dynamic routing algorithm, which enhances spatial relationship capture, and a tailored class weighting scheme optimized for the BraTS2020 dataset’s class imbalance. This pioneering evaluation on BraTS2020 demonstrates the hybrid model’s ability to preserve feature hierarchies and improve segmentation of irregularly shaped tumors. Figure 2 shows the illustrates the training and validation accuracy of the model over 25 epochs. Figure 3 illustrates the training and validation Accuracy of the model across 25 epochs.

**Fig 2.**
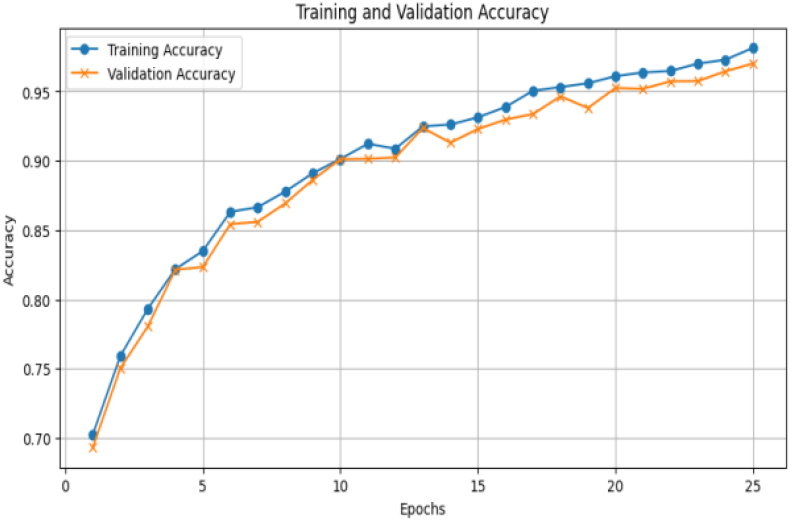
Training & Validation Accuracy

**Fig 3.**
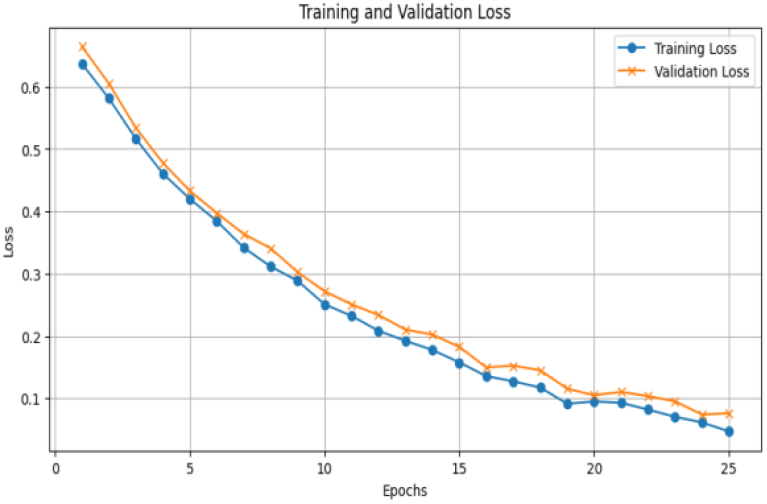
Training & Validation Loss

### 5.3 Observations

This section discusses the various observations.

#### 5.3.1 Class imbalance impact

Although class imbalance was addressed through class weighting, the results still showed slightly lower performance for the necrotic core class. This is likely because the necrotic core occupies a smaller area within the MRI scans, making it harder for the model to accurately identify and segment. However, the model still achieved an F1-score of 0.83 for this class, indicating a strong performance relative to its representation in the dataset.

#### 5.3.2 Generalization and overfitting

The use of data augmentation, early stopping, and reconstruction loss significantly reduced the risk of overfitting, as evidenced by the model’s close alignment between training and validation accuracy. These techniques encouraged the model to learn robust features that generalize well to unseen data. The high F1-score across all tumor classes in the test set further supports the model’s generalization capabilities.

#### 5.3.3 Computation efficeincy

Although Capsule Networks are computationally intensive, the use of GPU acceleration and mixed precision training allowed the model to train efficiently without sacrificing performance. The balance between computational complexity and performance was maintained, making the model viable for real-world applications in medical imaging.

## 6 Threats to validity

Primarily, the authors consider the class imbalance and computational limit issues as the major threats to the validity of the study.

### 6.1 Class imbalance

One of the most significant challenges in using CNNs for brain tumor detection is the class imbalance problem, where the majority of the dataset consists of normal brain tissue, while only a small portion represents tumor tissue. CNNs, like most machine learning models, tend to perform poorly in the presence of class imbalance, as they are biased toward the majority class (Buda et al., 2018; H. He & Garcia, 2009). In brain tumor datasets, non-tumor regions often outnumber tumor regions, which causes CNNs to be overly confident in classifying normal tissue, leading to a higher rate of false negatives. This is especially problematic in medical imaging, where detecting small or early-stage tumors is critical for timely treatment. To mitigate this issue, various techniques such as oversampling, undersampling, and class weighting are employed, but these techniques only offer partial solutions Table 1.1: Common Approaches to Address Class Imbalance

### 6.2 Computational limits

CNNs are computationally expensive, especially when applied to 3D medical imaging data like MRI scans. The large number of parameters in deep CNN architectures, such as U-Net and ResNet, require significant computational resources, making training and inference slow (Dou et al., 2017; Ronneberger et al., 2015). This complexity is a barrier to real-time applications, particularly in resource-constrained environments such as smaller hospitals or clinics

## 7 Conclusion and Future works

The study pioneered the use of a hybrid CapsNet + ResNet50 model with a custom 5-cycle dynamic routing algorithm for brain tumor detection and segmentation on the BraTS2020 dataset. This model, enhanced by a tailored class weighting scheme addressing class imbalance, achieved an overall accuracy of 98.4% with an F1-score of 0.87, outperforming traditional CNNs and benchmark architectures like U-Net. These results validate the effectiveness of the custom routing and weighting in preserving spatial information and enhancing segmentation accuracy, particularly for irregularly shaped tumors. Future work will explore refining the dynamic routing cycles and extending the weighting scheme to 3D MRI data.

## Declarations

## Data Availability Statement

The data utilized in this study is from the Brain Tumor Segmentation (BraTS2020) dataset, which is publicly available on Kaggle and was originally provided as part of the BraTS2020 competition. The dataset includes multimodal magnetic resonance imaging (MRI) scans and manual segmentation labels for brain tumor sub-regions. The scans are pre-processed to ensure consistency, including co-registration to a common anatomical template and interpolation to a uniform resolution. Researchers can access the dataset on Kaggle via the following link: BraTS2020 Dataset on Kaggle.

## Ethics Approval

Not applicable.

## Consent to Participate

All authors agreed to participate in this research.

## Consent for Publication

All authors have agreed to publish this article.

## Code, Data, and Materials Availability

The computer code supporting the findings of this study is openly available on Kaggle at the following permanent link: [res+caps_brain_tumor](https://www.kaggle.com/code/atsueric/res-caps-brain-tumor).

The dataset used in this research is the Brain Tumor Segmentation (BraTS2020) dataset, which is publicly accessible via Kaggle as part of the BraTS2020 challenge. The dataset can be accessed at: [BraTS2020 Dataset (Training + Validation)](https://www.kaggle.com/datasets/awsaf49/brats20-dataset-training-validation).

All materials, including preprocessing pipelines and model implementations, are available within the Kaggle notebook repository. No additional proprietary code, data, or materials were used in this study.

## Author Biographies

All authors contributed to the study’s conception and design. Data collection and preprocessing were performed by Paul Ammah, Experimental setup and setting up of the models were done by Eric K. A. Astu. First draft made by Solomon Mensah and Eric K. A. Atsu. All authors commented on previous versions of the manuscript. All authors read and approved the final manuscript.

